# Geriatrics 8 scores as a predictor of postoperative outcome in elderly patients with Head and Neck cancer in Rajavithi Supertertiary Care Hospital

**DOI:** 10.1101/2022.10.15.22281086

**Authors:** Sirikon Lertseree, Somjin Chindavijak

**Affiliations:** Center of Excellence in Otolaryngology Head & Neck Surgery, Rajavithi Hospital, Bangkok, Thailand

**Keywords:** Geriatric 8 score, Geriatric Head and neck cancer, post operative complication

## Abstract

**Background:** To determine preoperative Geriatric 8 (G8) score in predicting postoperative complications for elderly head and neck cancer patients.

**Material and Methods:** The prospective study in elderly head and neck cancer patients who underwent surgery during 30th January 2021-25th January 2022. G8 score were collected before surgery and analysed for the association with complication outcome.

**Result:** Of 104 patients included in this study, The mean age was 68.84 (SD =6.99 years). The Geriatric 8 (G8)score ≤ 14 which were frail group in 73 cases (70.2%) The Clavien-Dindo complications grade III-IV were 30 patients (28.8%). Among these groups, 26 patients (86.7%) was in frail groups and 4 patients (13.3%) with non frail group which is statistically significant different (p=0.019) and Odd ratio of 3.32, CI =1.01-10.87, p=0.048

**Conclusion:** The G8 score is a practical tool for prediction post operative complication in elderly Head and Neck Cancer surgery.

## Introduction

The new challenging for otolaryngologists is taking care of elderly Head Neck cancer patients. Every country in the world is experiencing the growth of elderly population whose age of 65 years and above that will reach over 1.5 billion by 2050^1^ In Thailand the old age population increase from 17.6% in 2018 to 18% in 2020^2^. The number of head and neck cancer also increase among this groups. Most of the standard treatment for Head Neck Cancer is major in surgery that concerned of postoperative complications in these elderly patients because of underlying disease and physiological change that affect the capacity of tolerability to the stress especially whose lesion was advanced stage. The chronological age alone is not a reliable predictor of postoperative complications as this cannot capture the physiologic heterogeneity prevalent in this population ^3,4^. Without the predictor tool, the plan of treatment trend to deviate from standard guideline due to patient factors of old age.

To identify the fit elderly for surgery or who need special care, the preoperative assessment tool is essential. The Traditional assessment that performed for every patients are concerned that was not sufficient to predict postoperative complications. Frailty is a term used for older adults and recognizes global limited reserve to withstand stressors. Frailty represents a state of reduced physiologic reserve associated with increased susceptibility to disability.^5^ By definition, a frail individual is highly susceptible to poor healthcare outcomes. The Comprehensive Geriatric assessment (CGA) Geriatric assessment for frailty were recommended by The Society for International Oncology in Geriatrics (SIOG) for administration to older patients who are receiving cancer care ^6^ The CGA was a mulitidimensional, multidisciplinary process which consistently included physical, functional, Co-morbidity, cognitive, nutrition, polypharmacy, social support and mental status, mobility/balance that was time consuming ^7,8^ Bellera et al^9^reported cut off value of 14 by Geriatric 8 (G8) score for elderly cancer patients who schedule for chemotherapy with sensitivity of 85% against the reference exam consisting of seven comprehensive geriatric assessment questionnaires with ease of application in practical work. The predictive value of the G8 score for postoperative complication of surgery were waiting for explored. Many studies reported of high accuracy for prediction of postoperative complications in other field of cancer treatment^10-,12^ but some reported of impaired G8 not be predictive tool ^13.^ And few reports in Head and Neck cancer surgery.

The objective is to study the application of preoperative G8 score whether or not be the predictive tool of postoperative complications in elderly head and neck cancer patients.

## Material and Method

This research was approved by the Ethics Committee of Rajavithi Hospital (EC number 63229). A prospective study was performed by collection of data from all patients who have been diagnosed with head and neck cancer with age of 60 years and over who had been treated by surgery at Center of Excellence in Otolaryngology Head and Neck surgery, Rajavithi Hospital during 30^th^ January 2021-25^th^ January 2022. All patients were evaluated by routine preoperative assessment which were CBC, LFT, electrolyte, BUN, Cr, EKG, Chest x-ray and asked for consent to evaluate of G-8 score by physicians in our department. The patients whose underlying or abnormal result of blood chemistry, EKG or chest x ray were sent for specialist consultation. All of these patients were pass of these evaluation and planned for operation. The data were recorded of age, sex, site of cancer, stage of cancer, type of operation include resection and reconstruction, intraoperative blood loss, duration of operation, post operative complications.

The G-8 score consisted of 8 components with total score from 0-17. These questionnaires were filled out at OPD which are : Component 1. Has food intake declined over the past 3 months due to loss of appetite, digestive problems, chewing, or swallowing difficulties? Response score of 0 = severe decrease in food intake, 1 = moderate decrease in food intake,2 = no decrease in food intake. Component 2. Weight loss during the last 3 months? Response score of 0 = weight loss >3 kg,1 = does not know,2 = weight loss between 1 and 3 kg, 3 = no weight loss. Component 3 : Mobility? Response score of 0 = bed or chair bound,1 = able to get out of bed/chair but does not go out,2 = goes out. Component 4 : Neuropsychological problems? Response score of 0 = severe dementia or depression1 = mild dementia, 2 = no psychological problems. Component 5 : BMI? _(weight in kg)/(height in m2)_ Response score of 0 = BMI <19, 1 = BMI 19 to <21,2 = BMI 21 to <23,3 = BMI ≥23. Component 6: Takes more than three prescription drugs per day? Response score of 0 = yes,1 = no. Component 7 : In comparison with other people of the same age, how does the patient consider his/her health status? Response score of 0.0 = not as good, 0.5 = does not know, 1.0 = as good, 2.0 = better. Component 8 : Age. Response score of 0: >85, 1: 80–85,2: <80. The G8 score ≤14 was defined as frailty patient.

The post-operative complications within 30 days were recorded and classify by Clavien-Dindo classification which were : Grade I as any deviation from the normal postoperative course without the need for pharmacological treatment or surgical, endoscopic and radiological interventions. Allowed therapeutic regimens are : drugs as antiemetics, antipyretics, analgesics, diuretics, electrolytes, and physiotherapy. This grade also included wound infections opened at the bedside. Grade II as requiring pharmacological treatment with drugs other than such allowed for grade I complications, Blood transfusions and total parenteral nutrition are also included. Grade III as requiring surgical, endoscopic or radiological intervention, Grade IV as Life threatening complication (including CNS complication) requiring IC/ICU management and Grade V as Death of patient.^14,15^.

The correlation of complications and each factors were studied included G8 score. Qualitative variables were described using counts and proportions. Quantitative continuous variables were described using means and standard deviations for normal data or medians and ranges otherwise. Associations between qualitative variables were tested using the chi-square test or the Fisher’s exact test. The Kruskal Wallis or Mann–Whitney U test was used for comparison of the continuous data and the Chi-square or Fisher’s exact test for categorical data. All tests were two-sided with a level of significance set at p < 0.05. Univariate and multivariate binomial logistic regression models were used to assess the Odds Ratio (OR) with 95% CI testing the ability of G8-based groups to predict perioperative morbidity after adjusting for all common preoperative covariates

## Result

There were 104 cases included, mean age was 68.84±6.99 years with males 71 cases (68.3%). History of underlying disease were 68 cases (65.4%) which were hypertension 46 cases (67.6%), Diabetes mellitus 21 cases (30.9%), Dyslipidemia 17 cases (25%), Pulmonary disease 12 cases (17.6%), cardiovascular diseases 5 cases (7.4%) and others 8 cases (11.8%).The Body Mass Index (BMI) overall mean 22.21 ± 4.3 kg/m^2^. The history of alcohol drinking and smoking found in 53 (51%) and 67(64.4%) respectively. The site of cancer were oral cancer 56 cases (53.8%), presented with advanced stage 86 cases (82.7%), the reconstruction of operation mainly were non-free flap in 97 cases (93.3%) and mean duration of operation was 5.31±2.41 hrs which more than 4 hrs 75 cases (72.1%). The America Society of Anesthesiologists score (ASA score) <3 in 78 cases (75%). The Geriatric 8 score (G8 score) ≤ 14 which defined as frail group in 73 cases (70.2%) as in Table 1.The number of cases in each categories of G8 score was that of 92 cases (88.5%) for age < 80 years, no decrease of food intake 59 cases (56.7%), moderate to severe food intake 44 cases (43.3%), can go out in mobility category of 97 cases (93.3%), no psychological problems in 104 cases (100%), Body Mass Index (BMI) ≥ 23 Kg/m2 in 41 case (39.4%), no history of taking medication more than 3 prescription drugs per day in 82 cases (78.8%) and health status as good compared with other people of the same age in 61 cases (58.7%) as in table 2

**Table 1.** Characteristic data (n=104)

| Characteristic | n (%) |
| --- | --- |
| **Gender** |  |
| Male | 71 (68.3) |
| Female | 33 (31.7) |
| **Age (Yrs), mean±SD** | 68.84±6.99 |
| 60-69 years | 61 (58.7) |
| 70-79 years | 31 (29.8) |
| ≥80 years | 12 (11.5) |
| **BMI (kg/m<sup>2</sup>), mean±SD** | 22.21±4.30 |
| < 18.5 kg/m<sup>2</sup> | 21 (20.2) |
| 18.50 – 22.99 kg/m<sup>2</sup> | 41 (39.4) |
| 23.00 – 24.99 kg/m<sup>2</sup> | 15 (14.4) |
| ≥ 25.00 kg/m<sup>2</sup> | 27 (26.0) |
| **Underlying disease** |  |
| No | 36 (34.6) |
| Yes | 68 (65.4) |
| Hypertension | 46 (67.6) |
| Diabetes Mellitus | 21 (30.9) |
| Hyperlipidemia | 17 (25.0) |
| Pulmonary disease | 12 (17.6) |
| Cardiovascular disease | 5 (7.4) |
| Others | 8 (11.8) |
| **Alcohol History** |  |
| No | 51 (49.0) |
| Yes | 53 (51.0) |
| **Smoking History** |  |
| No | 37 (35.6) |
| Yes | 67 (64.4) |
| **Cancer site** |  |
| Oral cancer | 56 (53.8) |
| Other Head Neck cancer | 48 (46.2) |
| **Cancer Stage** |  |
| Early stage | 18 (17.3) |
| Advanced stage | 86 (82.7) |
| **Reconstruction** |  |
| Free Flap | 7 (6.7) |
| Non-Free Flap | 97 (93.3) |
| **Duration of operation (hours)** |  |
| < 4 hours | 29 (27.9) |
| ≥ 4 hours | 75 (72.1) |
| **ASA score** |  |
| ASA score ≥ 3 | 26 (25.0) |
| ASA score < 3 | 78 (75.0) |
| **G8 score** |  |
| G8 score ≤14 | 73 (70.2) |
| G8 score >14 | 31 (29.8) |

BMI = Body Mass Index

**Table 2.** Descriptive of G8 Question Score (n=104)

| G8 Question | n (%) |
| --- | --- |
| **Age** |  |
| > 85 years | 5 (4.8) |
| 80 – 85 years | 7 (6.7) |
| < 80 years | 92 (88.5) |
| **Decrease food intake** |  |
| Severe decrease | 14 (13.5) |
| Moderate decrease | 31 (29.8) |
| No decrease | 59 (56.7) |
| **Weight loss** |  |
| Weight loss > 3 kg | 22 (21.2) |
| Does not know | 2 (1.9) |
| Weight loss between 1 – 3 kg | 34 (32.7) |
| No weight loss | 46 (44.2) |
| **Mobility** |  |
| Bed or chair bound | 0 (0.0) |
| Able to get out of bed/chair but<br>does not go out | 7 (6.7) |
| Goes out | 97 (93.3) |
| **Neuropsychological problem** |  |
| Severe dementia or depression | 0 (0.0) |
| Mild dementia | 0 (0.0) |
| No psychological problem | 104 (100.0) |
| **Body Mass Index (BMI)** |  |
| BMI < 19.00 kg/m<sup>2</sup> | 27 (26.0) |
| BMI 19.00 – 20.99 kg/m<sup>2</sup> | 16 (15.4) |
| BMI 21.00 to 22.99 kg/m<sup>2</sup> | 20 (19.2) |
| BMI ≥ 23.00 kg/m<sup>2</sup> | 41 (39.4) |
| **Takes more than 3 prescription drugs**<br>**per day** |  |
| Yes | 22 (21.2) |
| No | 82 (78.8) |
| Health status compare with other people of the same age |  |
| Not as good | 7 (6.7) |
| Does not know | 35 (33.7) |
| As good | 61 (58.7) |
| better | 1 (1.0) |

The overall major post operative complication were reported of Clavien-Dindo complication of Grade III-V in 30 cases (28.9%). Duration of operation and G8 score were statistically significant associated with complications groups but the history of smoking, alcohol drinking, site, stage and type of reconstruction were not associated as in table 3. Logistic regression analysis also demonstrated of statistically significant of postoperative complication by G8 score (OR =3.32, CI =1.01-10.87, p=0.048) as in table 4.

**Table 3.**
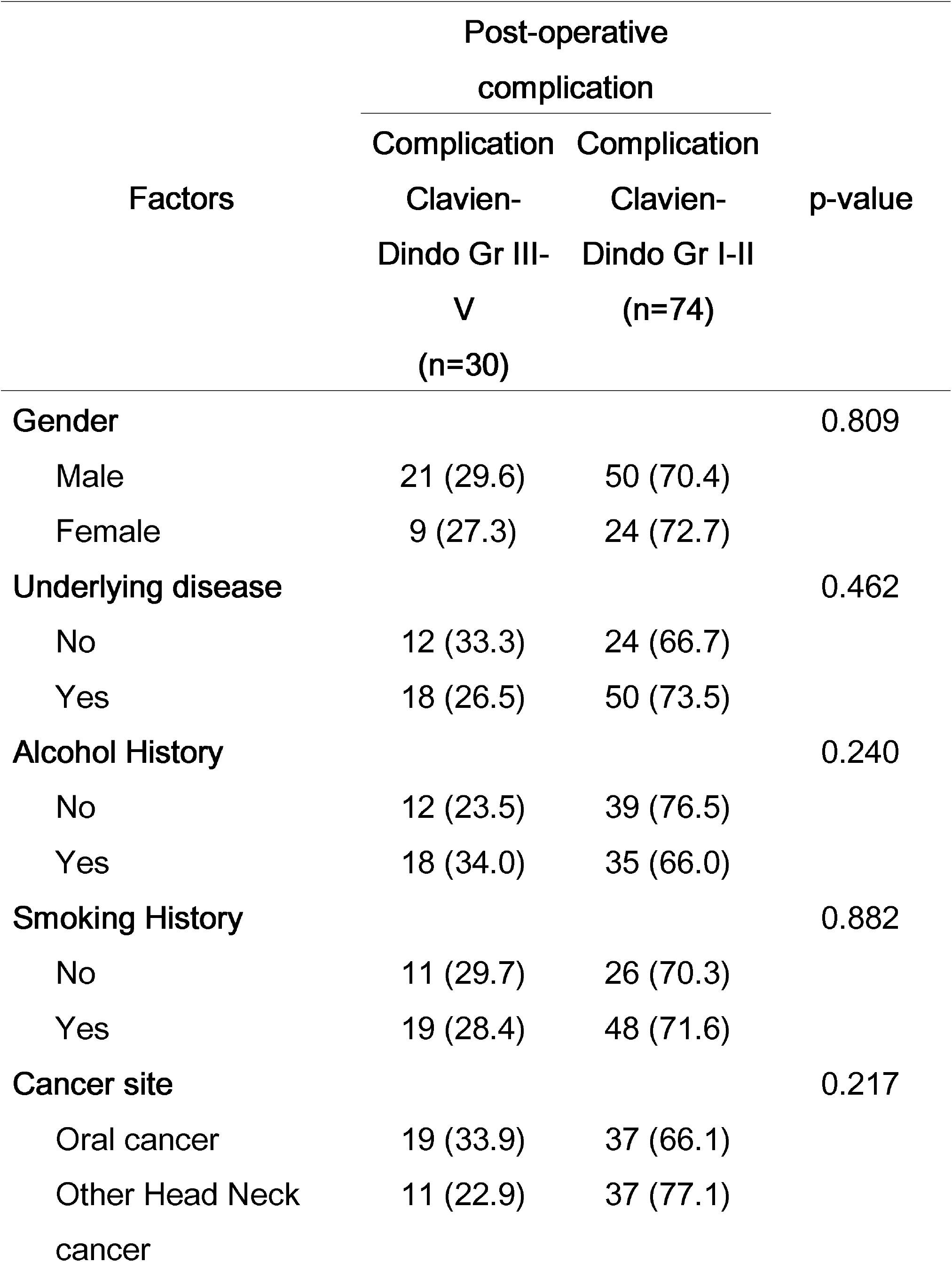
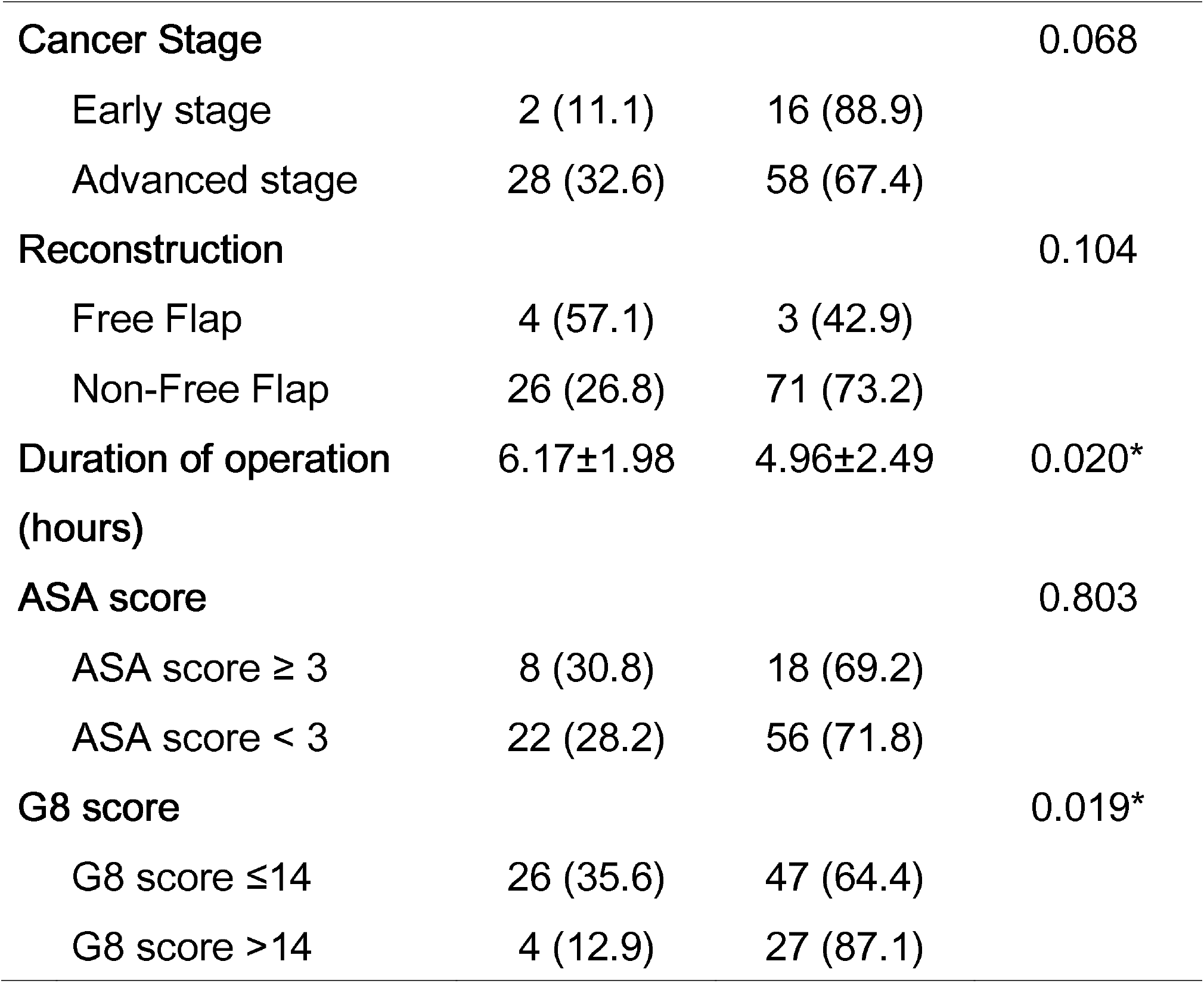
Factors associated with post-operative complication (n=104)

| Factors | Post-operative complication |  | p-value |
| --- | --- | --- | --- |
|  | Complication | Complication |  |
|  | Clavien-Dindo Gr III-V<br>(n=30) | Clavien-Dindo Gr I-II<br>(n=74) |  |
| **Gender** |  |  | 0.809 |
| Male | 21 (29.6) | 50 (70.4) |  |
| Female | 9 (27.3) | 24 (72.7) |  |
| **Underlying disease** |  |  | 0.462 |
| No | 12 (33.3) | 24 (66.7) |  |
| Yes | 18 (26.5) | 50 (73.5) |  |
| **Alcohol History** |  |  | 0.240 |
| No | 12 (23.5) | 39 (76.5) |  |
| Yes | 18 (34.0) | 35 (66.0) |  |
| **Smoking History** |  |  | 0.882 |
| No | 11 (29.7) | 26 (70.3) |  |
| Yes | 19 (28.4) | 48 (71.6) |  |
| **Cancer site** |  |  | 0.217 |
| Oral cancer | 19 (33.9) | 37 (66.1) |  |
| Other Head Neck cancer | 11 (22.9) | 37 (77.1) |  |
|  | Complication | Complication |  |
|  | Clavien-Dindo Gr III-V | Clavien-Dindo Gr I-II |  |
|  | (n=30) | (n=74) |  |
| **Cancer Stage** |  |  | 0.068 |
| Early stage | 2 (11.1) | 16 (88.9) |  |
| Advanced stage | 28 (32.6) | 58 (67.4) |  |
| **Reconstruction** |  |  | 0.104 |
| Free Flap | 4 (57.1) | 3 (42.9) |  |
| Non-Free Flap | 26 (26.8) | 71 (73.2) |  |
| **Duration of operation (hours)** | 6.17±1.98 | 4.96±2.49 | 0.020* |
| **ASA score** |  |  | 0.803 |
| ASA score ≥ 3 | 8 (30.8) | 18 (69.2) |  |
| ASA score < 3 | 22 (28.2) | 56 (71.8) |  |
| **G8 score** |  |  | 0.019* |
| G8 score ≤14 | 26 (35.6) | 47 (64.4) |  |
| G8 score >14 | 4 (12.9) | 27 (87.1) |  |

**Table 4.** Logistic Regression analysis for factors associated with post-operative complication

| Factors | Crude OR | p-value | Adjusted OR | p-value |
| --- | --- | --- | --- | --- |
| Duration of operation (hours) | 1.24 (1.02-1.50) | 0.027* | 1.20 (0.99-1.44) | 0.051 |
| G8 score |  |  |  |  |
| G8 score >14 | Ref |  | Ref |  |
| G8 score ≤14 | 3.73 (1.18-11.84) | 0.025* | 3.32 (1.01-10.87) | 0.048* |

## Discussion

Taking care of elderly head and neck cancer patients whose resection of primary cancer were planned is concerned due to increase in number of patients and high rates of postoperative complications. The preoperative assessment of elderly patients that fit for head and neck cancer surgery in our center was the same as the general population that scheduled for surgery. Nowadays many tools were reported for different assessment due to frailty in this group. The objective of assessment is to identify the elderly patients who were at risk of post operative complications and need intensive care or reconsider of management plan by surgery. All of these elderly patients pass the routine assessment but the complications rate were different among the groups. The comprehensive geriatric assessment (CGA)^16^ were accepted as the standard of care for evaluation the status of elderly patients which consisted of a set of six questionnaires:, Activities of Daily Living (ADL), Instrumental Activities of Daily Living (IADL), 4-item Geriatric Depression Scale (GDS-4), Mini Mental State Examination (MMSE), Mini Nutritional Assessment Short Form, and any falls in the previous year.The CGA was considered abnormal when a patient received an impaired score on at least one questionnaire. For practical usage, the CGA was time consuming but the G8 score takes 3-5 minutes and consists of seven item dealing with food intake, weight loss, mobility, neuropsychological problem, body mass index, prescription drug and self perception of health that more practical. ^17^

The main finding in the present study was the G8 score can predict the postoperative complications which is the practical point of new preoperative assessment for elderly patients. As the conventional assessment for the elderly patients who intended for schedule of head and neck surgery in our department who all were assessed as low risk for surgery but found as frail patients were 70.2% which higher than expected because these groups were already selected for surgery by traditional assessment Overall prevalence reported as Handforth et al that median prevalence of frailty across studies that identified frailty using CGA was 43% (range 7–68) ^18,19^ By the G8 score, the group of frailty were reported of higher complication rate than non-frailty in many studies ^20^ and included of free flap reconstruction in elderly head and neck cancer patients by Nagayama^21^ recently.

As our department is the supertertiary center that performed high volume of cases in major head and neck surgery per year The preoperative assessment tool that predict the postoperative outcome is important for making the care plan to prevent the postoperative complications in the elderly group which included the surgical and non surgical complications. The non surgical complications such as postoperative delirium, heart failure, myocardial infarction and pneumonia are usually less concern by Otolaryngologists due to the physical assessment such as normal EKG, normal blood chemistry and normal chest x-ray. There were few of Otolaryngology department in Thailand reported of G8 score for preoperative assessment. This report will guide the new assessment in elderly head and neck cancer patients.

The limitation of this study, there are a few of free flap reconstruction cases which is long duration operations and severe stress for elderly patients which concerned of postoperative complication whether G8 score can predict or not. The varieties of site of cancer and surgeons in this study may affected the complications that have to be controlled in the future study.

## Conclusion

The G8 score is one of the geriatric assessment tool that can apply in routine preoperative evaluation of elderly head and neck cancer patients for prediction of post operative complication and for plan of management in the frail group for decrease post operative morbidity and mortality.

## Data Availability

All data produced in the present work are contained n the manuscript

## Acknowledgement

I would like to thank all staff members at the Division of Medical Research, Rajavithi Hospital.

## Financial support

Rajavithi Hospital funding.

## Conflict of interest

none

## Notes

### Competing Interest Statement

The authors have declared no competing interest.

### Funding Statement

This study was funded by Rajavithi hospital

### Author Declarations

Ethic committee of Rajavithi Hospital gave EC 63229 approve for this work

